# Tracking national opinion about wastewater monitoring as a standard complement of public health tools in the United States

**DOI:** 10.1101/2023.06.16.23291485

**Authors:** A. Scott LaJoie, Rochelle H. Holm, Lauren B. Anderson, Heather D. Ness, Ted Smith

**Affiliations:** Department of Health Promotion and Behavioral Sciences, School of Public Health and Information Sciences, University of Louisville, 485 E. Gray St., Louisville, KY 40202, United States; Christina Lee Brown Envirome Institute, School of Medicine, University of Louisville, 302 E. Muhammad Ali Blvd., Louisville, KY 40202, United States

**Keywords:** community health, environmental monitoring, Privacy Attitudes Questionnaire, public opinion, sewer, wastewater-based epidemiology

## Abstract

National opinion on a wide variety of public health topics can change over time or have highly contextual nuisances. As the COVID-19 pandemic has progressed, we aimed to see how public perceptions have changed regarding the acceptance of using wastewater for community health monitoring in the metropolitan United States. This study is an annual update of prior inquiry into knowledge of wastewater-based epidemiology, privacy concerns surrounding the collection of wastewater samples and the use of data acquired along with privacy awareness from an online survey administered in the winter of 2023. We found public support for monitoring of toxins (90.8%), disease (90.6%), terror (86.8%), illicit drugs (70.5%), prescription medications (68.6%), and gun residue (59.8%) remained high. Most respondents supported monitoring of the entire city (77.6%). This longitudinal research has shown year-upon-year only slight increases in public support of wastewater monitoring for public health protection and continues to show an absence of significant nationwide privacy concerns as long as catchment population sizes ensure fully anonymous households. This support is conditional and is probably best understood through the lens of the COVID-19 pandemic where the value proposition of public protection was popularized. The survey also consistently showed the public would support expansion of wastewater monitoring as a standard complement of public health tools into other areas of public health protection.

## Introduction

The American public has opinions on a wide variety of health topics from smoking (Duch et al., 2018) to vaccination requirements (Haeder, 2021). In some cases, the public’s health awareness and beliefs change over time or are informed by highly contextual nuisances. One example is the increased public awareness of the harm smoking tobacco causes, such as lung cancer and the acceptance of scientific evidence that has evolved over the past 70 years (Duch et al., 2018). For smoking, the confluence of scientific evidence and public opinion is now enshrined in tobacco regulation. While wastewater monitoring for communicable and/or infectious human diseases has been a standard global public health tool for more two decades (Kilaru et al., 2023), the coronavirus disease 2019 (COVID-19) pandemic increased the use of this public health tool exponentially. However, the rate of change in the public’s awareness and acceptance of wastewater monitoring as a standard public health surveillance method as the pandemic has progressed have not been studied.

The intersection of ethics, a scientific or public health surveillance honor system, and public opinion for wastewater-based epidemiology (WBE) is complex. With current, widespread, and international pursuit of WBE technologies and methods, discussion of this intersection of science ethics and the public’s right to privacy is becoming more prominent in academic and policy arenas (Gable et al., 2020; Hrudey et al., 2021; National Academies of Sciences, Engineering, and Medicine, 2023). However, there have been few surveys on the United States national public opinion on this topic (Hill et al., 2022; Holm et al., 2022; LaJoie et al., 2022; Riback et al., 2023). There are four ethical principles in public health surveillance: common good, equity, respect for persons, and good governance (World Health Organization, 2017; Hrudey et al., 2021); these principals are not regulations or even guidance.

This study builds on previous research on public awareness and acceptance of WBE, privacy concerns regarding wastewater sample collection, and data usage, using an online survey administered in 2022 during the COVID-19 pandemic (LaJoie et al., 2022). Since the extent and severity of the pandemic changed from a year earlier, we aimed to investigate how public opinion may have changed regarding the awareness and acceptance of using wastewater for community health monitoring in the United States. Using the 2022 study as a baseline (Wave 1), we aimed to assess changing 2023 (Wave 2) opinions about WBE as a standard component of public health monitoring tools of the United States population in the period towards as the end of the national state of emergency neared.

## Materials and Methods

Our materials and methods are presented in detail in our earlier study (LaJoie et al., 2022). The recruitment matrix and data collection instrument were unchanged. In brief, adult research participants were recruited by Qualtrics XM (Provo, UT, USA) to complete a 15-min online survey about their knowledge and acceptance of WBE, perceptions of what is/should be monitored, where wastewater monitoring should occur, and support for, or opposition to, sewer monitoring as part of public health surveillance. The survey also included a privacy attitudes questionnaire (PAQ) (Chignell, Quan-Haase and Gwizdka J., 2003). The PAQ provides an indication of how people feel about their personal privacy under four domains, beyond sewer and public health, including: willingness to share personal information, exposure, willingness to be monitored, and interest in protection. Scores are calculated by summing the items within each domain (the number of items in domains differ, therefore, comparing one domain to another domain is not meaningful). The survey was limited to English-speaking, United States urban and suburban residents. Participants were compensated by Qualtrics XM. Responses for Wave 1 were collected between January and February 2022 and previously published (LaJoie et al., 2022). The updated data (Wave 2) presented in this study, were collected between February and March 2023.

### Data management and analysis

The Wave 2 results are reported, and changes between Wave 1 and Wave 2 are reported when the responses differed more than the margin of error (+/- 2% based on a sample size of 3,000 from an estimated United States population of 330 million with a confidence level equal to 95%). Margin of error is interpreted as the confidence interval around the sample proportion; in the present study, there is a 95% likelihood of the true population portion being contained within the margins of error. When the proportions do not differ between the two samples, only the Wave 2 response is reported. The responses support or strongly support were aggregated into one category. Data analysis was performed using the IBM Statistical Package for the Social Sciences (SPSS) software (version 28; Armonk, NY, USA).

### Ethics

The University of Louisville Institutional Review Board approved this project as Human Subjects Research (IRB number: 21.0877).

## Results

Wave 2 survey responses (n = 3,002) were collected between February 28 and March 15, 2023. The demographics of the current sample are comparable to Wave 1 collected in 2022 (n = 3,083), save for a higher percentage of Black, Indigenous, and People of Color in Wave 2 (21% versus 16%). The majority of respondents were female, white, older, college educated, and earning less than $40,000. Approximately 70% of the respondents described their living environment as mostly suburban and approximately 30% as urban. Residence in rural areas was an exclusion criterion.

### Awareness and Knowledge

Public health agencies routinely monitor various activities that may pose health threats, including testing for air and water pollution, restaurant inspections for food safety, and inspecting hotels, motels, and public pools. Awareness was highest regarding restaurant inspections (84.5%), hotel and motel inspections (76% in 2022 versus 72% in 2023), and monitoring of drinking water (66.5%) and public pools (63.4%) (Figure 1). Just under half of respondents were aware public health agencies monitor for air pollution (49.8%) and monitor sewers for public health threats (48%). The ranked order of highest to lowest of these six activities only had slight year-on-year changes. Awareness of WBE for public health was unchanged between Wave 1 and Wave 2.

**Figure 1.**
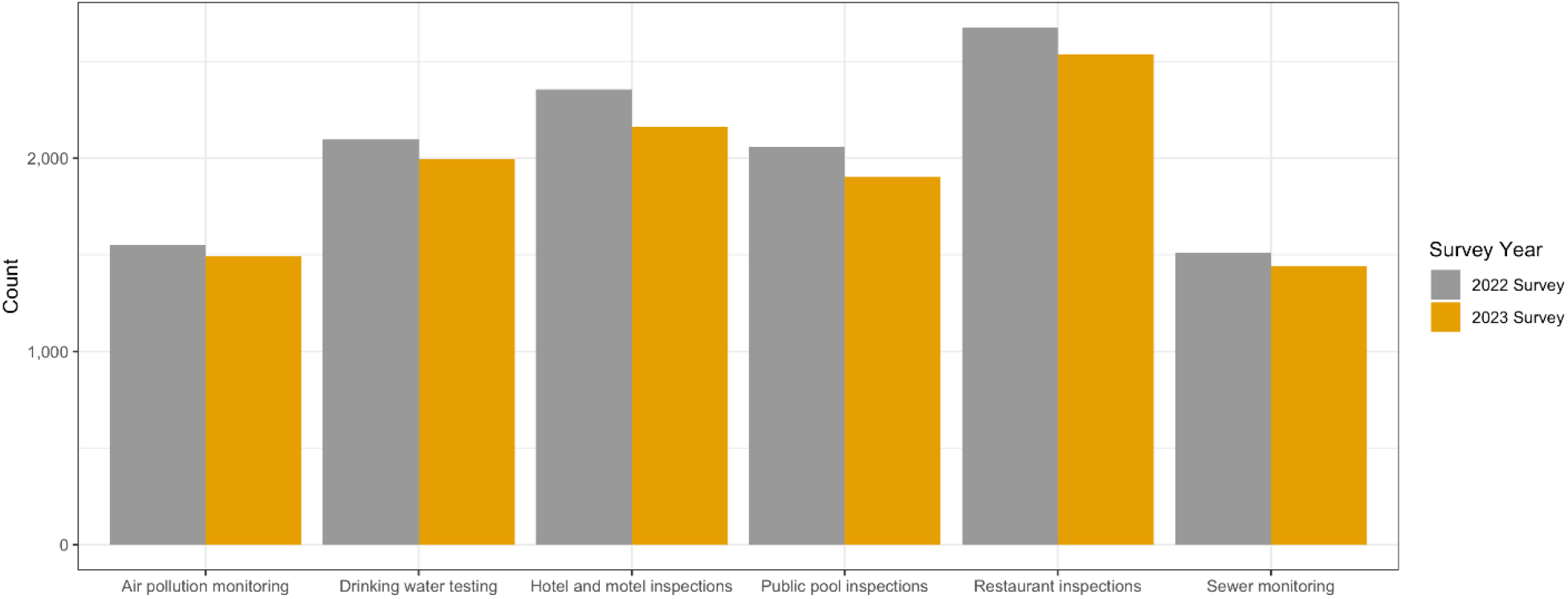
Frequency of awareness of public health monitoring activities, 2022 (LaJoie et al., 2022) and 2023 (this study) surveys.

Knowledge about how sewer monitoring could be used during a pandemic was measured with three items: could COVID-19 be detected in sewage (yes), could monitoring of sewer water determine which person or persons in a household had COVID-19 (no), and which of five stated assessment methods could most quickly detect COVID-19 in a community (sewer monitoring). Answers were summed to produce a knowledge score between 0 and 3 (all correct), with 17% correctly answering all three. Of note, 47% of survey respondents misbelieved sewer monitoring could determine which individual or individuals in a household has COVID-19.

### Support or Opposition to Monitoring

We presented 10 applications of wastewater monitoring (alcohol, disease, gun residue, healthy eating, illicit drugs, lifestyle behaviors, mental illness, prescription medicals, terroristic threats, and toxins) to participants and asked if they would support any of them. Support for the monitoring of each of the 10 applications increased or did not change from the 2022 survey (Figure 2). Areas that did not change in both surveys included support for monitoring of environmental toxins (e.g., industrial chemicals) (90.8%), deadly diseases (e.g., Ebola, Tuberculosis) (90.6%), terror (e.g., Anthrax) (86.8%), gun residue (e.g., bullet casings, gun powder) (59.8%) or alcohol (49%). Increases from year-to-year were seen in support for monitoring illicit drugs (from 67% to 70.5%), prescription medications (from 64.8% to 68.6%), mental illness (from 41.1% to 46.3%), healthy eating (from 31.2% to 37.1%), and for monitoring of lifestyle behaviors (e.g., smoking, birth control) (from 30.4% to 35.1%). Similar to the awareness question, the ranking of highest to lowest of these applications of wastewater monitoring priorities only had slight year-on-year changes showing consistent trends.

**Figure 2.**
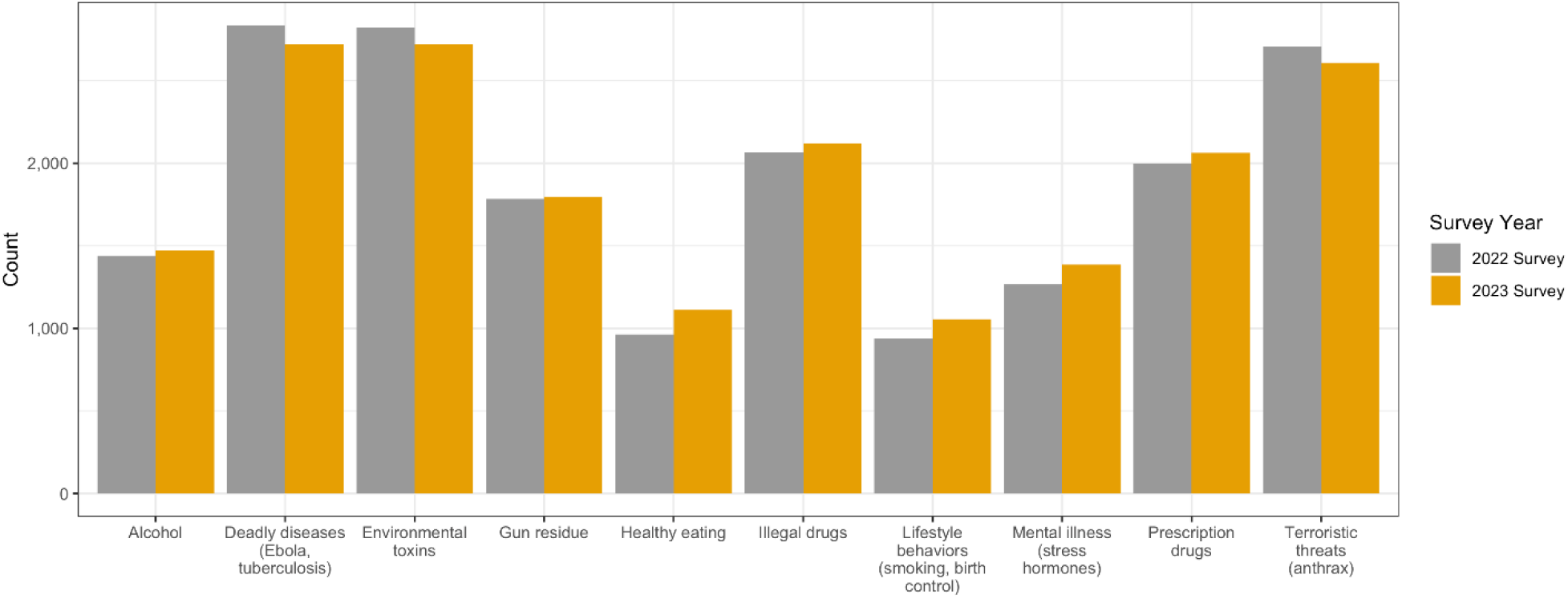
Frequency of support for 10 applications of wastewater monitoring, 2022 (LaJoie et al., 2022) and 2023 (this study).

In terms of support for monitoring areas or locations, only monitoring of the entire city (77.6%) was observed to not change from the 2022 survey (Figure 3). Responses were conditional upon having not endorsed “I would not support any monitoring of sewage water.” Increases in support were seen for monitoring areas of the city (from 20% to 22.3%), neighborhoods (23.3% to 26.7%), businesses (17.5% to 23.7%), prisons (20.3% to 23.7%), schools (25.4% to 28.6%) and houses (15.2% to 17.3%). There was a small decrease (from 11% to 9%) in those participants who would prohibit all sewer monitoring activities.

**Figure 3.**
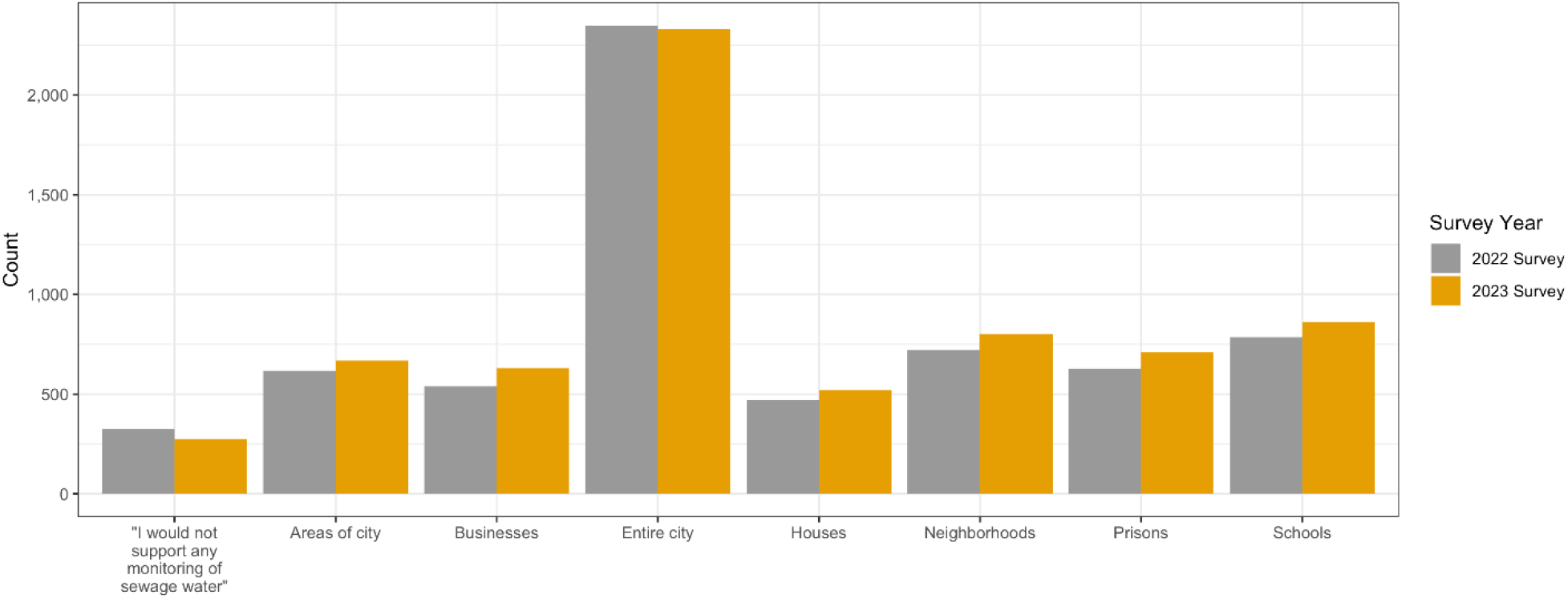
Scale of support for wastewater health monitoring activities in certain areas or locations, 2022 (LaJoie et al., 2022) and 2023 (this study). Responses are conditional upon having not endorsed “I would not support any monitoring of sewage water.”

### Privacy Concerns and Attitudes towards Privacy

When we asked about their level of confidence in city officials around keeping different types of information private, a majority of respondents were at least somewhat confident or highly confident. Rates of respondents indicating no confidence in city officials to keep different types of information private was unchanged from last year for lifestyle and behavioral (18.3%), financial information (17.5%) and health/medical information (15.9%). Generally, respondents trust their information will be handled appropriately.

The mean scores on three PAQ domains (exposure mean score 24.54, monitored mean score 30.05, and protection 31.39) did not differ from the 2022 survey, while the mean scores of personal information increased in 2023 to 24.00 from 23.61 (p<0.05). The increase in mean personal information score is interpreted as more comfort year-on-year in sharing information such as medical information with others.

## Discussion

These sequential data from a large nationwide sample, provide insights into public opinion about wastewater monitoring for use as a public health tool as the COVID-19 pandemic approached the end stage as a public health emergency. Other studies on the knowledge and acceptability of wastewater-based surveillance have been limited to a single city or state (Hill et al., 2022; Holm et al., 2022; Riback et al., 2023), and with only our prior nationwide study having been conducted during the Omicron variant peak (LaJoie et al., 2022). This survey was fielded when most pandemic-related restrictions had been lifted (though the federal emergency order had not yet expired) and shows even in a drastically different pandemic epoch, public opinion was unchanged in the support of the use of wastewater monitoring as a public health tool. In January 2023, the Pew Research Center (2023) found the coronavirus outbreak was one of the lowest American priorities (26%), while it had been among the top priorities in both 2021 and 2022; yet, defending against terrorism was reported as one of the highest 2023 American priorities (60%).

This is possibly most interesting, because select Category A biological terrorism agents, such as those requiring intensive public preparedness efforts, can be monitored in the environment (Sinclair et al., 2008), but this counter-terrorism activity is uncommonly conducted as part of regular surveillance by emergency management agencies. Also consistent, Ho et al. (2007) found that for public opinion on global health threats and infectious diseases, half were confident the federal government could deal with an avian flu outbreak and yet about 80% of Americans felt confident in a federal respond to anthrax health threats. Because WBE can monitor across a range of parameters, from infectious diseases to toxins, the recent interest around disaster and crisis response and preparedness has increased. Our survey results consistently showed public approval for wastewater use in surveillance of diseases, toxins, terrorist threats, and illegal activities, but individual health behavior surveillance is not acceptable. However, coordination and oversight for monitoring this wide range of activities is likely to be bifurcated and siloed.

For example: public health agencies are likely to oversee disease data and pandemic preparedness while environmental agencies would coordinate toxin data and security and law enforcement agencies would oversee terrorist and illegal activity wastewater data. Inter-agency wastewater monitoring coordination and scope in line with public preference requires further consideration when a range of parameters can be analyzed from a single sample aliquot.

Survey results around the sampling scale, in which privacy of anonymous wastewater sampling is conducted and protected, are consistent from 2022 to 2023. Our survey results indicate a boundary exists, particularly around, household scale monitoring. The average American household is 2.5 people (U.S. Census Bureau, 2022). Together, this indicates that a wastewater sample catchment scale be large enough to contain more than 3 positive cases of the wastewater monitoring parameter of interest. In the case of rare health threats and infectious diseases this may result in large wastewater sample catchment areas to avoid the potential stigmatization of the population being sampled. There are also ethical issues around quantifying personal information, such as human genome reads possible in gut metagenome sequencing data (Tomofuji et al., 2023). Although this is technically feasible with WBE, current wastewater monitoring precedent operates on the honor system that these samples are not to be used for individual human genetic monitoring.

As the pandemic approached the end of the public health emergency designation and hospitalization and deaths were declining in February 2023, our Wave 2 survey results indicated public awareness was unchanged, but support increased. Even though a lot has changed over the last year, what has not changed is that in this same time period, mentions of “wastewater” and WBE related terms in popular media remained relatively high (Figure 4; Cision, 2023). Previous public opinion research (Ho et al., 2007) found public attention paid to news peaked during global health threats and infectious diseases and wain in the period towards the outbreak tail. No evidence was found in our study to support the hypothesis that after a year of continued media coverage of sewer monitoring, respondents were more aware of, and more knowledgeable about, sewer monitoring for the detection of COVID-19.

**Figure 4.**
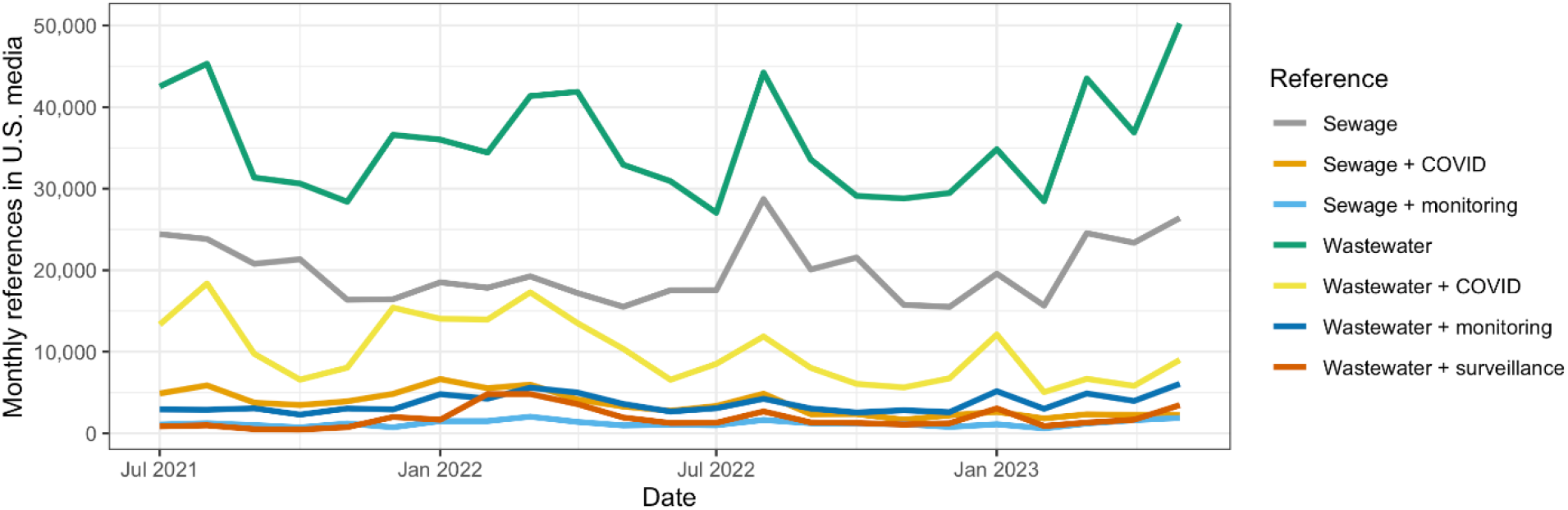
United States media mentions of “wastewater” and related terms, July 2021–May 2023 (Cision, 2023).

This study has several limitations. Outside the United States, public opinion may be different on this subject. Additionally, our invitation to participate was limited to the urban and suburban population and excludes the rural community.

Through this national survey of public opinion, we identified four key themes which could inform public policy for the long-term integration of wastewater monitoring in the public health arena:

1. **Support is conditional**. Sampling is most supported when catchment population sizes ensure fully anonymous households. Further, the targets of monitoring, such as pathogens, toxins, biological and chemical terrorist threats and illegal activities are generally considered acceptable.
2. **There is an overall concern about anonymity**. Where scientifically possible, our survey inferred from the overall concern about anonymity that there should be a transition from the honor system to guiding policy that wastewater monitoring not be used for individual human genetic monitoring.
3. **Wastewater monitoring is supported at levels comparable with other public health surveillance activities**. Like pool and restaurant inspections, health departments should set standard complement policy and funding for wastewater monitoring targets and priorities.
4. **Support is beyond global health threats and infectious diseases**. Toxins and terrorist threats offer an opportunity for wastewater surveillance expansion with high levels of public support.

## Conclusion

This longitudinal research survey has shown slight increases in public support of wastewater monitoring for public health protection and continues to show an absence of significant nationwide privacy concerns as long as catchment population sizes ensure fully anonymous households. This support is conditional and is probably best understood through the lens of the COVID-19 pandemic where WBE’s value towards public protection was popularized. However, the survey also consistently showed the public would support expansion of wastewater monitoring as a standard complement of public health tools into other areas of public health protection.

## Data Availability

All data produced in the present study are available upon reasonable request to the authors.

## Data Availability Statement

The data underlying the results presented in the study are available as a supplement.

## Author Contributions

Conceptualization: Ted Smith.

Formal analysis: A. Scott LaJoie, Rochelle H. Holm.

Methodology: Ted Smith.

Project administration: Ted Smith.

Supervision: Ted Smith.

Writing – original draft: Rochelle H. Holm.

Writing – review & editing: A. Scott LaJoie, Rochelle H. Holm, Lauren B. Anderson, Heather D. Ness, Ted Smith.

### Funding

This work was supported by grants from the James Graham Brown Foundation and the Owsley Brown II Family Foundation. The funders had no role in the study design, data collection and analysis, decision to publish, or preparation of the manuscript.

